# Subclade K influenza A (H3N2) viruses display partial immune escape with preserved cross-neutralisation in a UK population

**DOI:** 10.64898/2026.01.27.26344933

**Authors:** Kieran Dee, Ryan M Imrie, Laura Mojsiejczuk, Savitha Raveendran, Hanting Chen, Gerardo Guio Archila, Mosab S Aljeri, Verena Schultz, Ziping Wang, Sarah K Walsh, Junsen Zhang, Edward Hutchinson, Brian J Willett, David L Robertson, Joseph Hughes, Oscar A MacLean, Joe Grove, Emma C Thomson, Christopher J R Illingworth, Pablo R Murcia

## Abstract

We examined whether the recent emergence of influenza A(H3N2) subclade K, associated with an unusually early influenza season in the Northern hemisphere, was accompanied by a reduction in human population immunity. Using virus neutralisation assays on pre-epidemic human sera collected in May 2025, we found evidence of moderate antigenic drift. Further, vaccines used in the 2024/2025 season induced cross-neutralising immunity. These findings provide timely insight for interpreting recent influenza epidemiology and informing vaccine strain selection.

## MAIN TEXT

The unusually early start of influenza A(H3N2) activity in the UK in 2025^1^ raised concerns about the emergence of a highly mutated virus with reduced population immunity. Subclade K of influenza A(H3N2) (IAV-K), first detected in the UK in July 2025 (GISAID accession EPI_ISL_20152713), harbours multiple mutations in the haemagglutinin (HA) relative to other subclade J viruses, including the A/Thailand/8/2022-like viruses used in vaccines during the 2024/2025 season (Figure 1A–B).

**Figure. 1.**
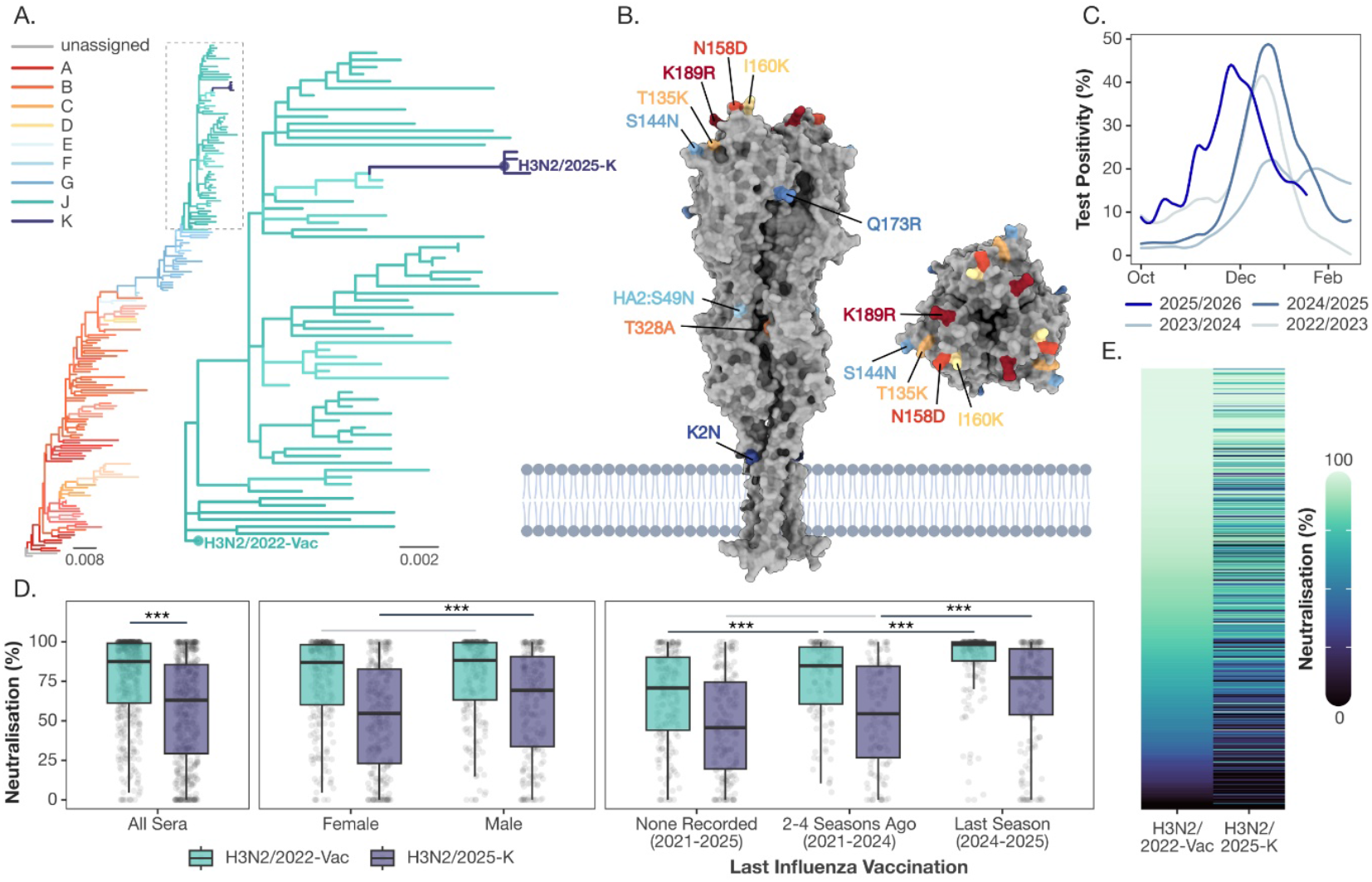
**A**. Evolutionary history of seasonal influenza A/H3N2 haemagglutinin sequences belonging to clade 3C and descendant subclades, compiled from public databases (data available up to January 20^th^, 2026). Tips are coloured by subclade assignment (see key). A zoomed-in view highlights the phylogenetic relationship between the vaccine strain (H3N2/2022-Vac) and the circulating subclade K representative (H3N2/2025-K). Branch lengths are drawn to scale (substitutions per site). **B**. Protein structure model of IAV-K HA trimer. Surface representation of the H3N2 IAV-K HA trimer (side profile, left; top-down view, right) predicted using AlphaFold3. Lineage-defining mutations are labelled and colour-coded. Labels are mapped to a single HA protomer, with the exception of the buried T328A mutation which is shown on an adjacent protomer. Note that while glycans are not rendered, T135K and S144N represent the loss and gain of *N*-linked glycosylation sites, respectively. A schematic representation of the viral lipid membrane is shown for the side view. **C**. Laboratory confirmed influenza cases in Scotland from the 2022/2023 season until January 12^th^ of the 2025/2026 season. **D**. Neutralising activity of serum samples against recombinant influenza viruses harbouring the surface glycoproteins of H3N2/2025-K, and H3N2/2022-Vac. **E**. Heat map showing the percentage neutralisation value for all samples used in serological assays. Each row represents an individual sample, arranged in order of H3N2/2022-Vac neutralisation, and columns show the virus tested.

IAV-K was associated with an early onset (Figure 1C) and a delayed end of the influenza season in the Northern and Southern^2^ hemispheres, respectively. As immune escape variants are associated with seasonal epidemics^3^, we investigated whether the accumulation of mutations in the IAV-K HA generated a reduction in population immunity. Neutralising activity of 486 serum samples derived from the NHS Greater Glasgow & Clyde adult patient population collected in May 2025 were evaluated against IAVs bearing the surface glycoproteins of both A/England/01837755/2025 (H3N2/2025-K), and A/Thailand/8/2022 (H3N2/2022-Vac, the vaccine strain deployed in the 2024/2025 season). Note, May 2025 was chosen as this is the end of the Northern hemisphere 2024/2025 influenza season, and before the detection of IAV-K in the UK.

Figure 1D shows that H3N2/2025-K neutralisation was on average 20% lower than for H3N2/2022-Vac (SE = 1.7%, p < 0.001) after accounting for differences between sex, age group, and vaccination history. For H3N2/2022-Vac, more recent vaccination was associated with increased neutralising activity: sera from individuals vaccinated during the previous influenza season (2024/2025) showed ∼9% higher neutralisation than those last vaccinated between the 2021-2024 seasons (SE = 2.1%, p < 0.01), while individuals last vaccinated in the 2021-2024 seasons displayed ∼9% increased neutralisation relative to those with no vaccination recorded between 2020-2025 (SE = 2.7%, p < 0.001). For H3N2/2025-K, neutralisation showed a weaker association with vaccination history, with little evidence of a difference between individuals with no vaccination recorded between 2020–2025 and those last vaccinated in the 2021–2024 seasons (difference ∼4%, SE = 3.5%, p = 0.49). In contrast, individuals vaccinated in the 2024–2025 season showed ∼10% higher neutralisation than those vaccinated between 2021–2024 (SE = 3.4%, p < 0.01). H3N2/2022-Vac and H3N2/2025-K neutralisation across sera was positively correlated (Figure 1E), with H3N2/2022-Vac neutralisation explaining ∼53% of the observed variation in H3N2/2025-K neutralisation (logit-scale R^2^ = 0.53, p < 0.0001). Sera from males showed a modest (∼7%) but significant increase in neutralisation against H3N2/2025-K (SE = 2.6 %, p < 0.01) but not H3N2/2022-Vac (difference ∼2%, SE = 1.8%, p = 0.28).

In sera from a large UK adult population, we show evidence of antigenic drift in subclade K IAV(H3N2). While reduced haemagglutination inhibition to IAV-K has been reported using ferret antisera^3^, our findings measure pre-epidemic human population immunity directly.

Our data are consistent with the recent epidemiology of IAV(H3N2), in which subclade K viruses were associated with an early seasonal peak but did not sustain prolonged transmission. While moderate antigenic drift was sufficient to alter epidemic timing, preserved cross-reactive immunity, particularly among recently vaccinated individuals, may have constrained epidemic duration.

## Acknowledgements

We gratefully acknowledge all data contributors (i.e., the authors and their originating laboratories responsible for obtaining the specimens, and their submitting laboratories for generating the genetic sequence and metadata and sharing via the GISAID Initiative.

## METHODS

### Ethics

Ethical approval for the collection of residual samples from Clinical Biochemistry was provided by NHSGGC Biorepository (application 837).

### Serum samples

Serum samples were provided by the National Health Service Greater Glasgow and Clyde (NHS GG&C) Biorepository. Random residual biochemistry serum samples (n = 486) from primary (general practices) and secondary (hospitals) health care settings were collected by the NHSGGC Biorepository. Associated metadata, including age, sex, date of sample collection, and date of influenza vaccination were retrieved from the NHS GG&C Safe Haven.

### Viruses

Viruses were rescued by reverse genetics as previously described^4^. Viruses carried the internal genomic segments of A/Puerto Rico/8/34 (PR8) and the surface glycoproteins (HA and NA) of A/England/01837755/2025 (H3N2/2025-K, GISAID accession EPI_ISL_20210731) or A/Thailand/8/2022 (H3N2/2022-Vac, GISAID accession EPI_ISL_14991375) Plasmids encoding the internal genomic segments of PR8 were a kind gift of Prof. Ron Fouchier^5^. Plasmids encoding the HA and NA of H3N2/2022-Vac and H3N2/2025-K were synthesised and cloned by GeneArt Thermo Fisher.

### Neutralisation assays

MDCK-SIAT1 cells were seeded at a density of 3 x10^5^ cells/ml ∼24h before infection. For the initial screen, sera were diluted 1:75 in DMEM with 0.25% bovine serum albumin (BSA). Then 60 µl of infectious medium containing 1.2 x10^3^ plaque forming units (PFU) of either H3N2/2025-K or H3N2/2022-Vac was added to 60 µl of each serum dilution in triplicate. The mixtures were incubated at 37°C for 1 h and 100 µl was added to the cells. The input control consisted of 60 µl of virus being added to 60 µl of DMEM 0.25% BSA. Polyclonal anti-sera raised in sheep against the HA of A/Cambodia/e0826360/2020(National Institute for Biological Standards and Controls: 21/118) was used as a positive neutralization control. Plates were incubated at 37°C, 5 % CO_2_ for 16 h and then fixed in 10% formalin. Cells were permeabilized with 1% Triton-X for 10 min. Virus positive cells were detected using a monoclonal antibody raised against IAV nucleoprotein (NP) (European Veterinary Labs, Clone EBS-I-238) at a dilution of 1: 1500 and then a secondary antibody (goat anti-mouse IgG Alexa Flour 488, A-11001) also at 1: 1500. Positive cells were counted using a Nexcelom Celigo imaging cytometer. For each plate, the mean total count for the input control was calculated. The total virus-positive cell count for each sample was then expressed as a percentage of the mean input count. These values were subtracted from 100, to give a percentage neutralization value.

### Statistical analysis

Neutralisation data was analysed as proportions using beta regression, adjusted using a Smithson-Verkuilen transformation to avoid exact values of 0 or 1. A forward model building approach was used, beginning with a null (intercept only) structure and assuming constant precision. Plausible explanatory variables – virus (H3N2/2025-K, H3N2/2022-Vac); vaccination recency (last vaccinated in the 2024-2025 influenza season, last vaccinated in the 2021-2024 influenza seasons, no record of vaccination across the 2021-2025 influenza seasons); age group (18-49, 50-64, 65+); sex; and experimental block – were evaluated sequentially. Model extensions were retained if Akaike information criterion (AIC) indicated sufficient support for the increased model complexity. Additions to the precision model were considered before additions to the fixed-effects model, and first order effects were evaluated before second- and then third-order interaction terms. This resulted in a precision model with first-order effects of virus and experimental block, and a fixed-effects model with all possible first-order effects and two second-order interactions: one between virus and vaccination recency, and another between virus and sex. Effects of individual explanatory variables are reported as estimated marginal means on the response scale, averaged over the levels of other covariates with equal weighting, and the statistical significance of pairwise comparisons was assessed using asymptotic z-tests. Correlations in neutralisation between H3N2/2025-K and H3N2/2022-Vac were assessed using Pearson correlation coefficients calculated on logit-transformed neutralisation values. All analyses were performed in R version 4.5.2 using the betareg (v3.2.4) and emmeans (v1.11.1) packages.

### Phylogenetic analysis

Influenza A virus sequences were retrieved from the NCBI Entrez databases using the taxonomic identifier and an in-house Python tool interfacing with the E-utilities API. This dataset was further enriched with sequences from GISAID, including all records available as of 20 January 2026. Analysis was restricted to HA coding sequences. Records failing basic quality control were excluded (sequences with excessive ambiguity, atypical genetic divergence, HA length < 1680 nucleotides, or inconsistent host annotations). All duplicate records were removed. To reduce dataset size while preserving diversity, human-derived sequences were selected and then clustered with MMseqs2 (v15.6f452) using a 0.988 sequence identity threshold (--cluster-mode 2 --cov-mode 1 --min-seq-id 0.988). Representative sequences, together with clade 3C references and the viruses of interest, were aligned using MAFFT (v7.453) with default parameters. A maximum-likelihood phylogeny was inferred with IQ-TREE (v2.4.0) under the best-fit substitution model. The subtree corresponding to clade 3C and descendant lineages was selected and visualised in RStudio using ggtree (v3.16.3). Subclade assignments (A–K) used to annotate and colour the tree were obtained with Nextclade (v3.18.1).

### Molecular modelling

The IAV-K HA trimer structure was predicted using a local installation of AlphaFold3^6^ (inference performed on an NVIDIA A800 GPU), achieving an average pLDDT prediction confidence of 84.2. To stabilise the orientation of the transmembrane domains, 25 molecules of oleic acid were included in the prediction. Visualisations were generated in ChimeraX^7^; the N-terminal signal peptide and oleic acid molecules were omitted from the final figures for clarity.

### Influenza surveillance data

Weekly laboratory-confirmed influenza virus infections from individuals presenting to General Practitioners with respiratory symptoms between October 3^rd^, 2022 and January 12^th^, 2026, were obtained from the Public Health Scotland (PHS) Community Acute Respiratory Infection (CARI) surveillance dataset.

## Data availability

All data used in this study can be found on GitHub: https://github.com/ryanmimrie/Publications-2026-H3N2-K-Cross-Neutralisation

